# Costs and predictors of early readmissions in patients with Infective Endocarditis. Utilizing the Nationwide Readmission Database

**DOI:** 10.1101/2021.06.04.21258024

**Authors:** Jesan Zaman, Amod Amritphale, Christopher Malozzi, Nupur Amritphale, Mukul Sehgal, Omar Bassam

## Abstract

**BACKGROUND:** There have been previous studies detailing the variables involved in readmissions in patients with a primary admission diagnosis of infective endocarditis – however those studies were done prior to the 2015 change in AHA guidelines and introduction to ICD-10 codes.

**OBJECTIVES:** The aim of this study was to describe the frequency, causes, factors, and costs associated with infective endocarditis encounters.

**METHODS:** Utilizing the 2017 national readmission database (NRD), we identified all patients that were admitted with infective endocarditis. These patients were evaluated for the rates, predictors, and costs of unplanned 30 days readmissions. Weighted analysis was performed to obtain nationally representative data.

**RESULTS:** 56,357 patients were identified to have been admitted with a diagnosis of infective endocarditis of whom 13,004 patients (23%) were readmitted within 30 days of the index discharge. The most common causes of readmission were septicemia (15.1%), endocarditis and endocardial disease (10.5%), heart failure (9.5%), and complication of cardiovascular device, implant or graft, initial encounter (5.6%). Data showed that there were certain comorbidities that resulted in a higher risk of being readmitted, these include chronic kidney disease, COPD, tobacco use, and hepatic failure. Cost of readmissions per patient was approximately $22,059 (IQR $11,630 - $49,964).

**CONCLUSIONS:** Thirty-day unplanned readmissions remain a significant issue affecting nearly 1 in 6 patients with infective endocarditis. This is associated with significant mortality and financial burden. Multi-disciplinary approach may help decrease readmissions, reduce complications, and improve overall outcomes as well as the overall quality of life of our patients.

## Introduction

Despite a low incidence rate of 3-10 individuals per 100 person-years, infection of the endocardium and/or heart valves, known as Infective Endocarditis (IE) is a potentially lethal process associated with significant mortality and morbidity^(1)^.

The mortality rate of IE is alarmingly high as detailed in a prior study with in-hospital rates of up to 20% and a 1-year mortality rate of up to 40%^(1)^. In addition, patients with IE were found to be at a higher risk for unplanned readmissions, a study by Aggarwal et al (2020) conducted over 6 years period, consisting of 187,438 patients with a primary diagnosis of IE, showed 24% of patients were readmitted within 30 days. Over half of the re-admissions (51%) occurred within the first 11 days of discharge^(2)^. Various factors have been implicated to achieve a better prognosis for IE patients ranging from early diagnosis and initiation of treatment, to different combinations or medical therapy, and surgical intervention during index admission; however, they require reevaluation to represent current practice patterns due to recent changes in the IE guidelines in relation to diagnosis, antimicrobial therapy, and management of complications^(3)^.

With the advent of ICD-10, the complexity of diagnosis/coding has increased multi-fold, which provides greater specificity to the research and more ability to pinpoint factors that may alter management^(4)^. This in addition to updated practice patterns with updated society guidelines, we evaluated large real-world data from the Nationwide Readmissions Database (NRD), to understand comorbidities and predictors associated with early unplanned readmissions in patients discharged with IE.

We followed the direction of previous well conceived studies using the database that have evaluated the risks of readmissions in patients with carotid stenting and pediatric sepsis^20, 21^.

## Methods

The NRD is a national representative sample of all-age, all-payer discharges from U.S. nonfederal hospitals which comes from the Agency of Healthcare Research and Quality. It contains data from 28 geographically dispersed states across the U.S. It has about 18 million discharges from the year 2017 (Weighted estimated to roughly 36 million discharges). To be fair however we have presented a flowchart in Figure 3 that gives the unweighted numbers. The NRD holds full information on both primary hospitalization and following readmissions within the states. In this study we utilized the data that represents 60% of the U.S. population and 58.2% of all U.S. hospitalizations.

**Figure 1:**
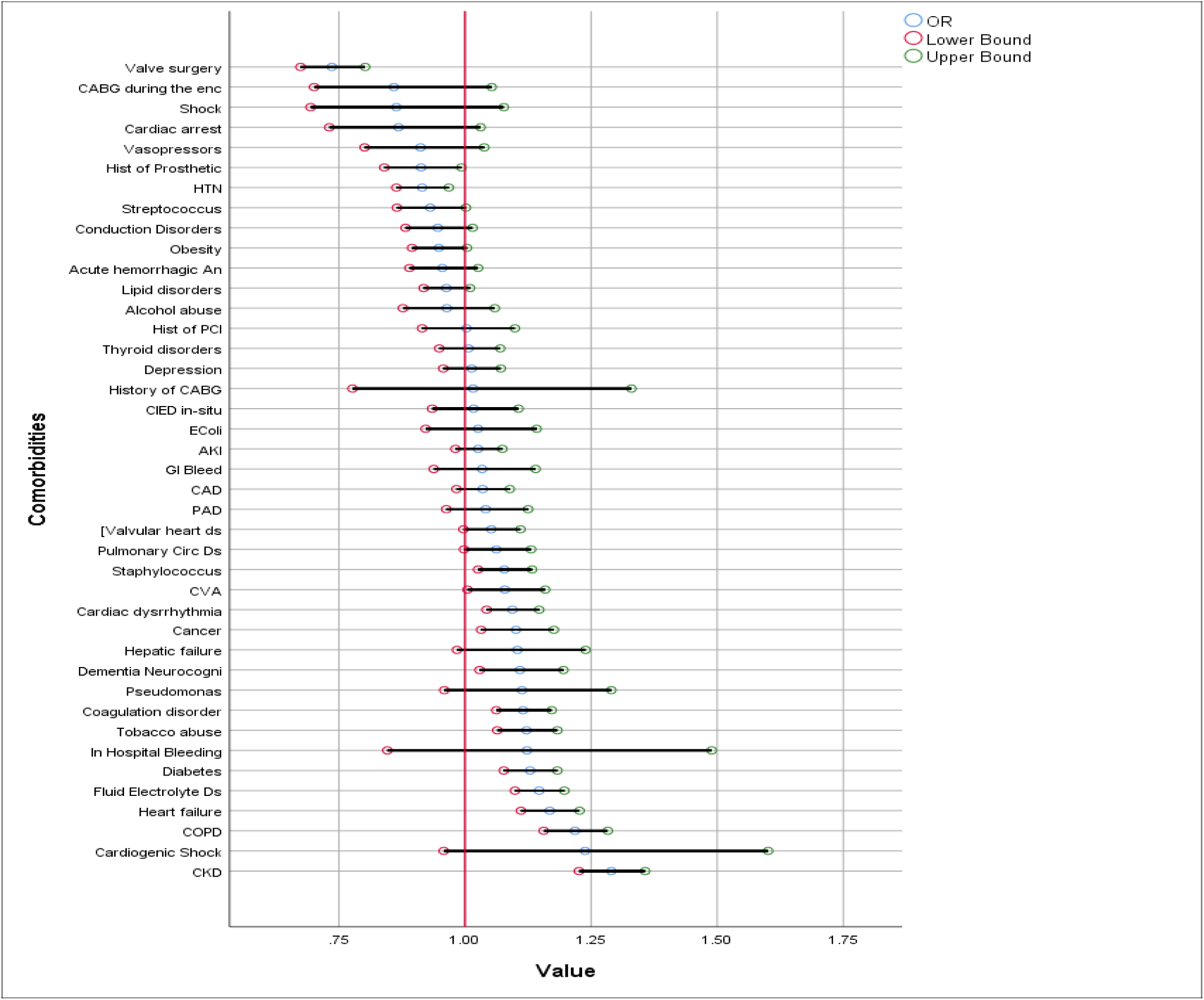
Multivariable logistic regression analysis – Forrest Plot. **Methods:** Utilizing the data retrieved, we created a Forrest plot to better visualize the differences in odds ratios of variables **Results:** Patients with Chronic Kidney Disease (CKD) showed to have the highest odds at 1.29 (1.22-1.36) of readmission while those who received valve surgery showed to be a protective factor at decreased odds at 0.83 (0.76-0.91). **Discussion:** Our study utilizing the NRD data from 2017 and ICD10 codes showed that CKD was a comorbidity that we should be paying more attention to during admissions and closer follow up may help in decrease readmission odds. Also, taking a closer look to re-evaluate surgical options earlier in admission may help in prognosis and decrease admission rates as well.

**Figure 2:**
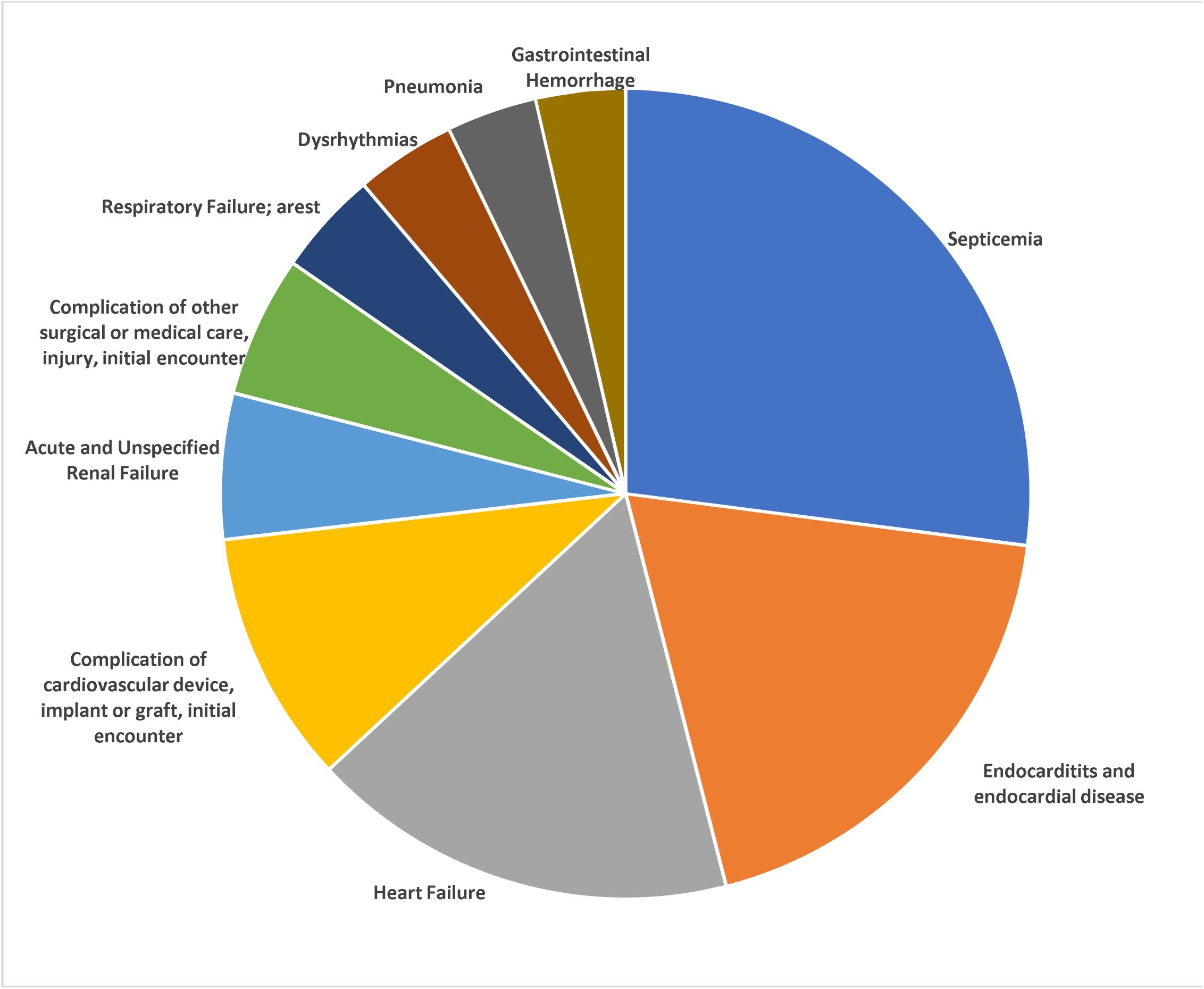
Top 10 frequencies of primary diagnosis category for readmissions encounters. **Methods:** Utilizing the data retrieved, we created a pie chart to better visualize comorbidities that were most present in readmission encounters **Results:** Patients discharged after an initial admission due to infective endocarditis were more likely to be readmitted due to septicemia (13.8%). **Discussion:** Infective endocarditis can easily lead to septicemia in patients, therefore it would be beneficial to repeat labs for markers of inflammation with closer follow up or re-evaluate duration of antibiotic therapy during the initial admission.

**Figure 3:**
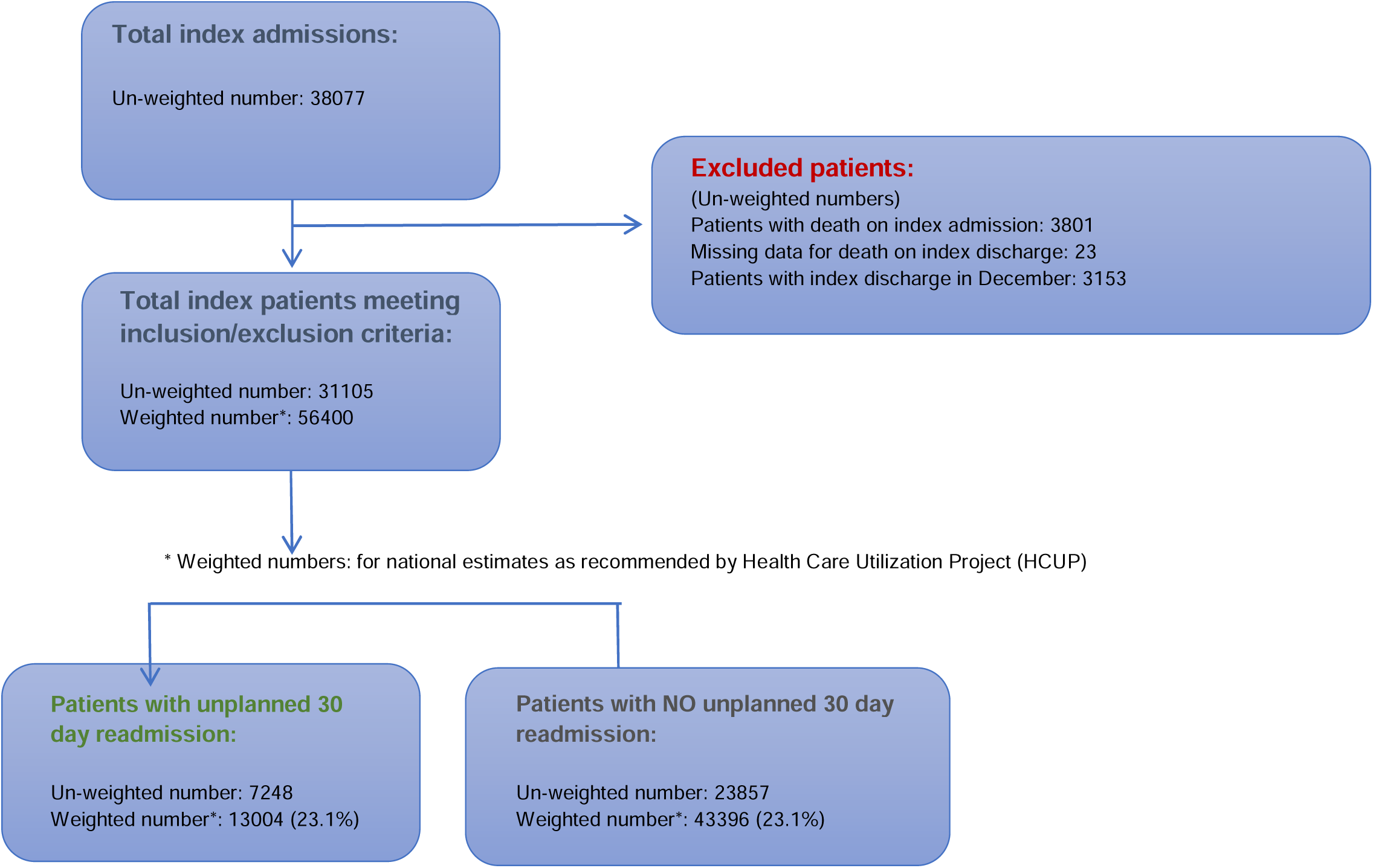
Flowchart of Unweighted Numbers.

Within 2017, hospitalizations and rehospitalizations were determined using a de-identified unique patient linkage number assigned to each patient, which let us track them across hospitals within a state. Each patient was assigned up-to 40 diagnosis codes and 25 procedure codes for each hospitalization. Often times a diagnosis established when a patient, presents is based on initial symptoms and then the overall main diagnosis of infective endocarditis is subsequently discovered during the actual course of admission. There is no way to tell which was the initial diagnosis and thus up to 40 diagnosis codes were utilized in our study. Additionally, patient information is note carried on from year to year and so it is counted as a new patient/diagnosis; patients cannot be tracked across years. Patients that had elective admissions were also not counted as an unplanned readmission. We also wanted to ensure that regarding readmission it was not just a short 1-night admission to ensure follow up, and this is taken care of because the NRD only captures patients that have inpatient status, not observation which is less than 24hrs. The University of South Alabama Institutional Review Board (IRB) approved the study for an exempt status.

We defined infective endocarditis with the procedure codes from ICD-10 as seen in Appendix Online Table 1. The primary outcome was first unplanned readmission after being diagnosed with infective endocarditis during the year 2017. Even if a patient had multiple such admissions in a year, we only used the first encounter for the analysis. Patients admitted in December were excluded as they may not have 30 days of follow-up, leading to immortal time bias. In addition, any patient that died during the initial hospitalization or had missing data were excluded.

**Table 1:**
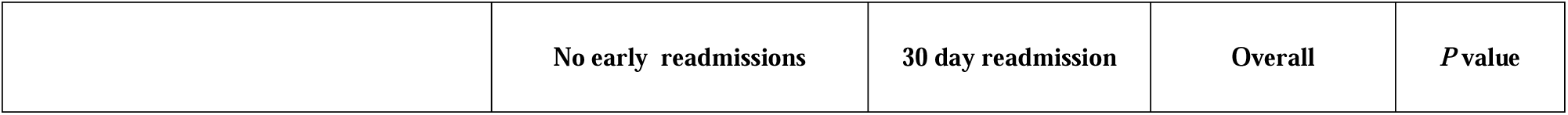
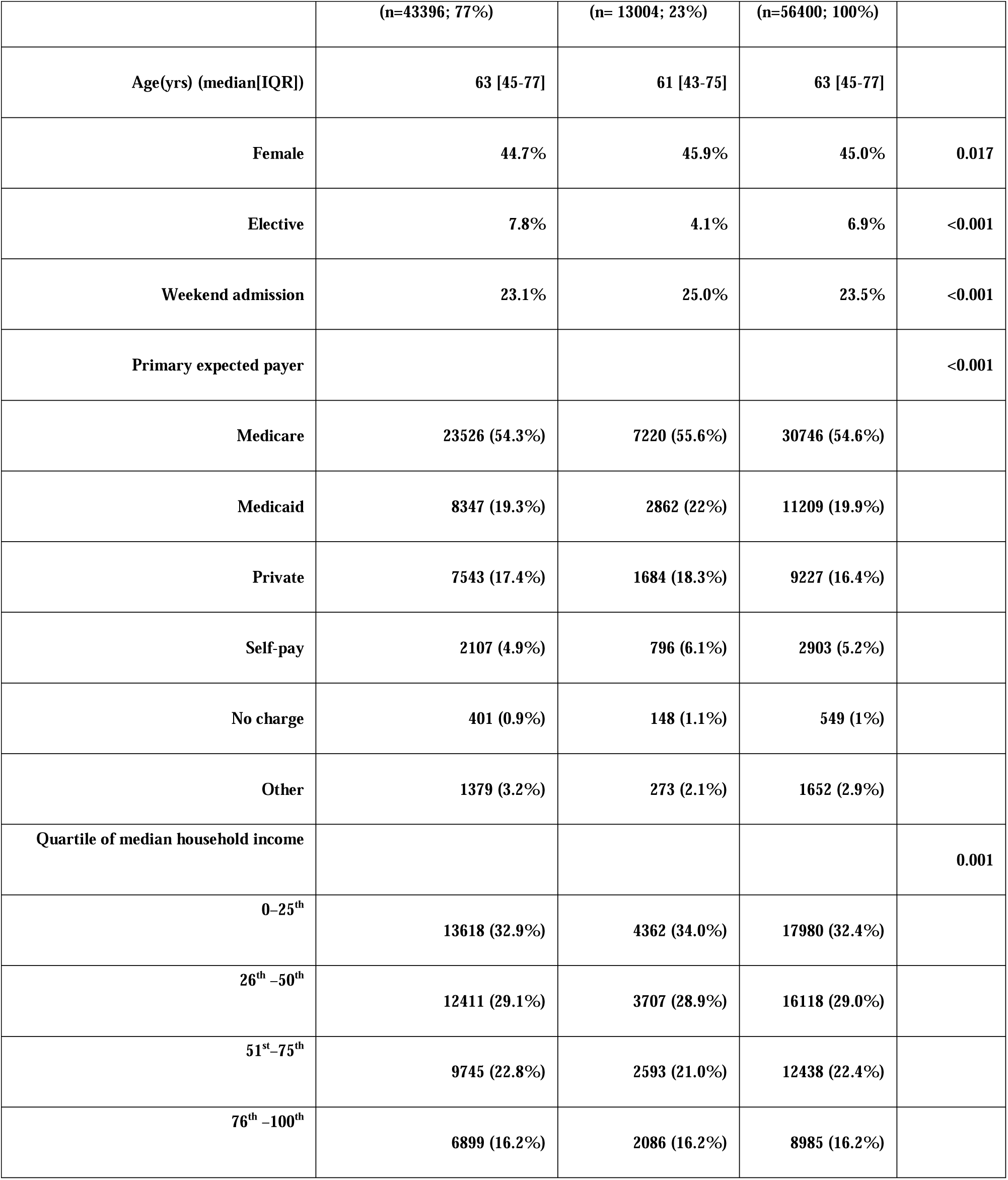
Baseline characteristics during index admission for Infective endocarditis.

|  | No early readmissions | 30 day readmission | Overall | P value |
| --- | --- | --- | --- | --- |

|  | (n=43396; 77%) | (n= 13004; 23%) | (n=56400; 100%) |  |
| --- | --- | --- | --- | --- |
| Age(yrs) (median[IQR]) | 63 [45-77] | 61 [43-75] | 63 [45-77] |  |
| Female | 44.7% | 45.9% | 45.0% | 0.017 |
| Elective | 7.8% | 4.1% | 6.9% | <0.001 |
| Weekend admission | 23.1% | 25.0% | 23.5% | <0.001 |
| Primary expected payer |  |  |  | <0.001 |
| Medicare | 23526 (54.3%) | 7220 (55.6%) | 30746 (54.6%) |  |
| Medicaid | 8347 (19.3%) | 2862 (22%) | 11209 (19.9%) |  |
| Private | 7543 (17.4%) | 1684 (18.3%) | 9227 (16.4%) |  |
| Self-pay | 2107 (4.9%) | 796 (6.1%) | 2903 (5.2%) |  |
| No charge | 401 (0.9%) | 148 (1.1%) | 549 (1%) |  |
| Other | 1379 (3.2%) | 273 (2.1%) | 1652 (2.9%) |  |
| Quartile of median household income |  |  |  | 0.001 |
| 0–25<sup>th</sup> | 13618 (32.9%) | 4362 (34.0%) | 17980 (32.4%) |  |
| 26<sup>th</sup>–50<sup>th</sup> | 12411 (29.1%) | 3707 (28.9%) | 16118 (29.0%) |  |
| 51<sup>st</sup>–75<sup>th</sup> | 9745 (22.8%) | 2593 (21.0%) | 12438 (22.4%) |  |
| 76<sup>th</sup>–100<sup>th</sup> | 6899 (16.2%) | 2086 (16.2%) | 8985 (16.2%) |  |

ICD-10 codes were used to define pertinent clinical variables. Data on status of hospital being teaching or non-teaching, urban or rural, the control/ownership, and even hospital bed sizes were collected. We performed a cost analysis by analyzing data on index admission length of stay (days), index admission cost (U.S.$), discharge destination to home/self-care, transfer to other hospital, care home, discharge against medical advice, on primary expected payer, quartile of median household income, readmission length of stay (days), readmission charges & costs (U.S.$), and readmission death. We multiplied the hospital charges with the Agency for Healthcare Research and Quality’s all-payer cost-to-charge ratios to determine costs. The first diagnosis on the basis of Clinical Classification Software codes (CCSR) determined the causes of readmission. All ICD-10 and CCSR codes used in this study are presented in the appendix section.

Statistical analysis was performed using IBM SPSS Statistics for Windows, version 1.0.0.118 (IBM Corp., Armonk, N.Y., USA). We used 2-sided tests with a significance level of 0.05 for statistical analysis. We examined baseline characteristics of participants and tested for statistical differences using the Pearson Chi Square test for categorical variables and Mann-Whitney U-Test for continuous variables with no readmission as the reference group. To determine the predictors of readmission within each time period with adjustments for all variables except the elective variable we utilized multivariable logistic regressions.

## Results

### Population Characteristics and Descriptive Results

30,413,839 patient encounters from January through December 2017 among hospitalized patients 18 years of age and older were included in the NRD. We identified those patients diagnosed with infective endocarditis using a weighted analysis. After index admission went infective endocarditis, 13,004 (23%) patients had an unplanned 30-day readmission.

General population and hospital data can be found in Tables 1 and 2. The median length of days of readmission was 10 days (IQR 5 to 18). 9.2% of the total and admission population received valvular surgery and among those readmitted, 7.7% received valvular surgery. The most common pathogen on readmission was Staphylococcus with 25.4%, followed by streptococcus which made up about 8%. Compared with the patients who were not readmitted within 30 days, those readmitted were more likely to have comorbidities chronic kidney disease, COPD, and tobacco use disorder. Additionally, those who received valvular surgery (95.7% of patients that got valve surgery received it during index admission) were less likely to have been readmitted according to our data with a odds ratio of 0.73 (0.67-0.80, P<0.001). Infective endocarditis is associated with significant morbidity and mortality and Table 3 shows some of those values we obtained in our study.

**Table 2:** Hospital, Discharge, and Length of Stay Data from Index Admissions of Infective Endocarditis.

|  | No early readmissions | 30 day readmission | Overall |
| --- | --- | --- | --- |

|  | (n=46343; 82.2%) | (n= 10048; 17.8%) | (n=56391) | *P* value |
| --- | --- | --- | --- | --- |
| **Hospital bed size** |  |  |  | **<0.001** |
| Small | 4533 (10.4%) | 1423 (10.9%) | 5956 (10.6%) |  |
| Medium | 31604 (72.8%) | 9454 (72.7%) | 41058 (72.8%) |  |
| Large | 7260 (16.7%) | 2127 (16.4%) | 9387 (16.6%) |  |
| **Control/Ownership of Hospital** |  |  |  | **0.202** |
| Government, nonfederal | 4533 (10.4%) | 1423 (10.9%) | 5956 (10.6%) |  |
| Private, not-profit | 31604 (72.8%) | 9454 (72.7%) | 41058 (72.8%) |  |
| Private, invest-own | 7760 (16.7%) | 2127 (16.4%) | 9387 (16.6%) |  |
| **Hospital Urban Rural Designation** |  |  |  | **<0.001** |
| Large metropolitan areas with at least 1 million residents | 23591 (54.4%) | 7693 (59.2%) | 31284 (55.5%) |  |
| Small metropolitan areas with less than 1 million residents | 16272 (37.5%) | 4553 (35.0%) | 20825 (36.9%) |  |
| Micropolitan areas | 2671 (6.2%) | 587 (4.5%) | 3258 (5.8%) |  |
| Not metropolitan or micropolitan (non-urban residual) | 862 (2.0%) | 171 (1.3%) | 1033 (1.8%) |  |
| **Hospital teaching status** |  |  |  | **<0.001** |
| Metropolitan non-teaching hospital | 9102 (21.0%) | 2685 (20.6%) | 11787 (20.9%) |  |
| Metropolitan teaching hospital | 30761 (70.9%) | 9560 (73.5%) | 40321 (71.5%) |  |
| Non-metropolitan hospital | 3533 (8.1%) | 758 (5.8%) | 4291 (7.6%) |  |
| **Discharge destination** |  |  |  | **<0.001** |
| **Home/self-care** | **14762 (34.0%)** | **3474 (26.7%)** | **18236 (32.3%)** |  |
| **Home health care** | **10300 (23.7%)** | **2927 (22.5%)** | **13227 (23.5%)** |  |
| **Discharge against medical advice** | **1848 (4.3%)** | **1522 (11.7%)** | **3370 (6%)** |  |
| **Length of stay and cost analysis** |  |  |  |  |
| **Index admission length of stay(days)**<br>**(median[IQR])** | **9 (5-18)** | **10 (5-18)** | **9 (5-18)** | **<0.001** |
| **Index admission cost (U.S.\$)**<br>**(median[IQR])** | **\$20241 (\$10,280 - \$45,184)** | **\$22059 (\$11630 - \$45964)** | **\$20642 (\$10609 - \$45381)** | **<0.001** |
| **Re-admission length of stay(days)**<br>**(median[IQR])** |  | **10 (5-18)** |  |  |
| **Re-admission cost (U.S.\$)**<br>**(median[IQR])** | | **\$22059 (\$11630 - \$45964)** | | |

**Table 3:**
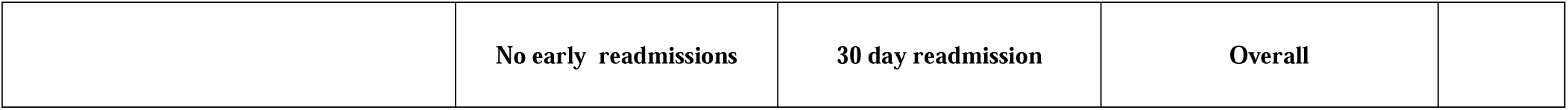
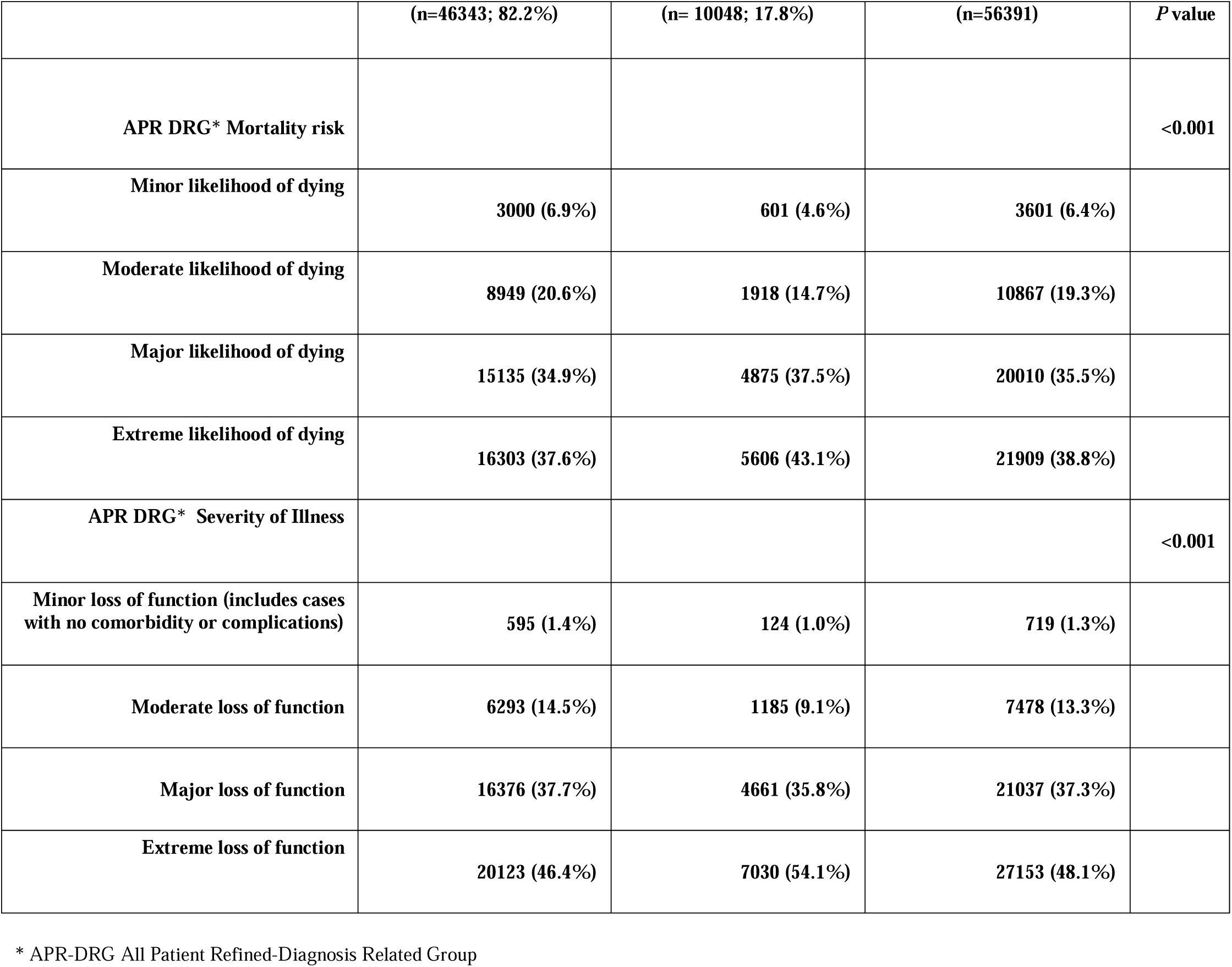
Morbidity and Mortality Data of Index Admissions from Infective Endocarditis.

### Cost analysis and length of stay results

The median hospital costs of index encounters were $20,241 [IQR $10,280 – $45,184]. The median length of stay for index admission and readmission encounters were 9 days (IQR 5-18 days) and 10 days (IQR 5-18 days, *P*<0.001) respectively.

## Discussion

Due to a change in infective endocarditis guidelines in 2015 we decided to reassess the quality of hospital care and performance utilizing 30 day readmissions in the year of 2017 as a metric. In this study we found that unplanned 30 day readmission rate after a primary diagnosis of infective endocarditis was 23%. This was relatively similar to the rates after primary diagnosis of cardiac dysrhythmias (16.73%), Urinary tract infections (18.31%), and acute myocardial infarction (19.66%). The condition with the highest percentage of readmissions was congestive heart failure at 28.05%^(5)^. A previous study analyzing patients with infective endocarditis after discharge, reported slightly higher rates of readmissions at about 24.8%^(1)^. This indicates a slight positive trend in treatment that may have occurred due to the change in guidelines in 2015, however readmission statistics are still a concern, and risk factors should be evaluated to decrease the rate further, to alleviate hospital costs. Studies have also shown that most readmissions occurred within the period immediately after discharge, and nearly 1 in 12 patients died during those events which further emphasize a need to focus on early prevention and intervention on the risk factors and comorbidities that were highlighted in our study^(1)^. Table 4 details the comorbidities and procedure related factors during index admissions for infective endocarditis.

**Table 4:**
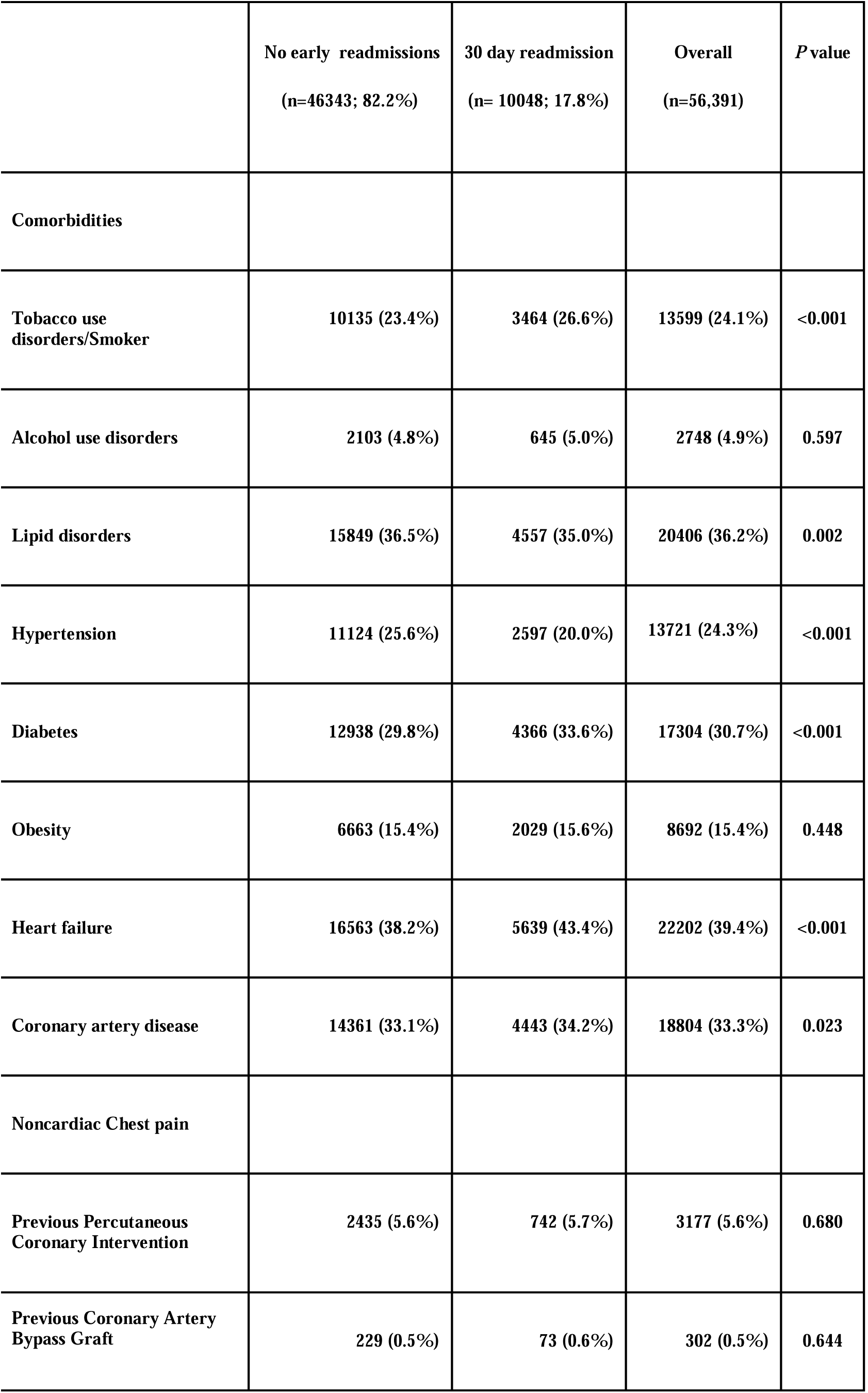
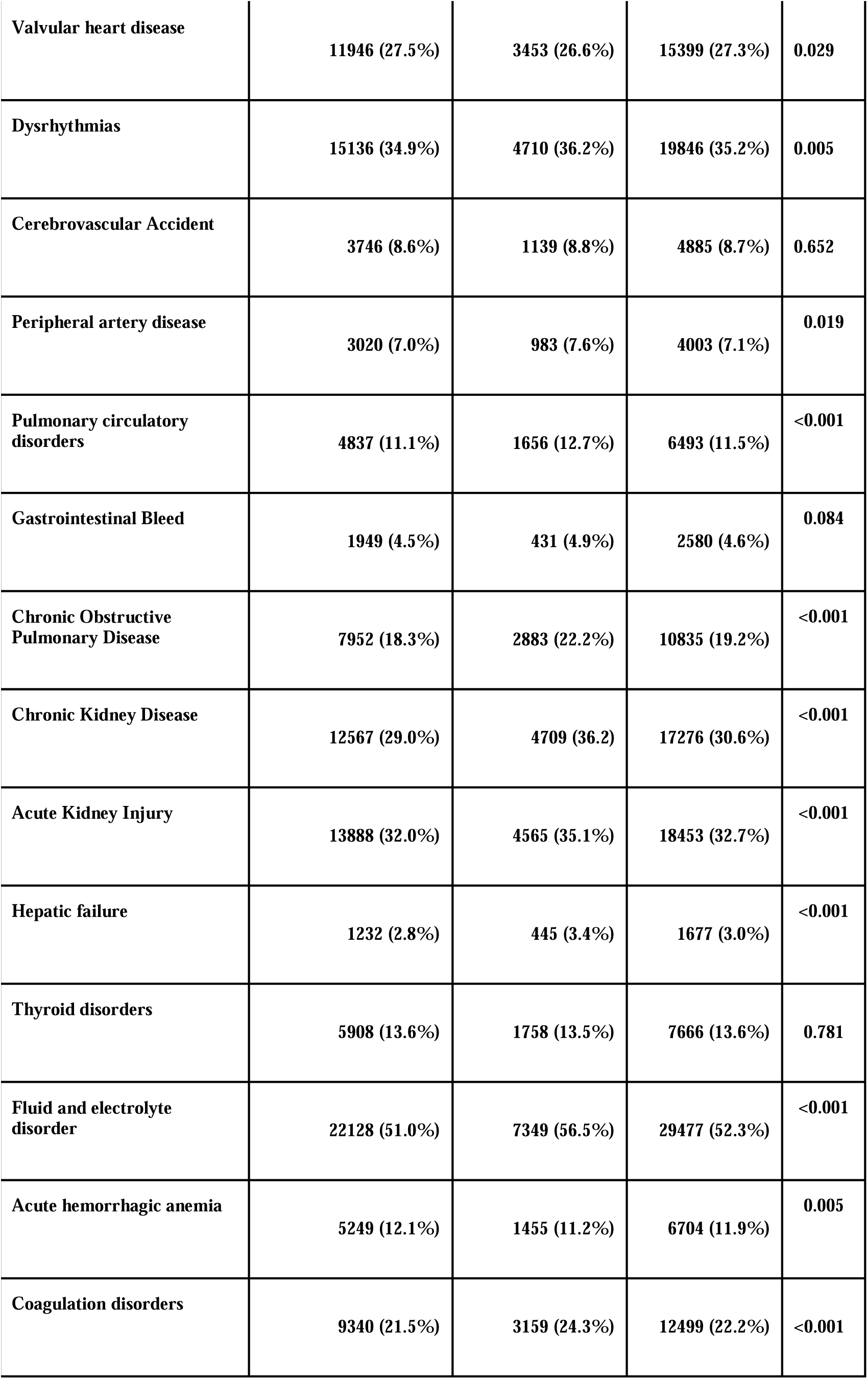
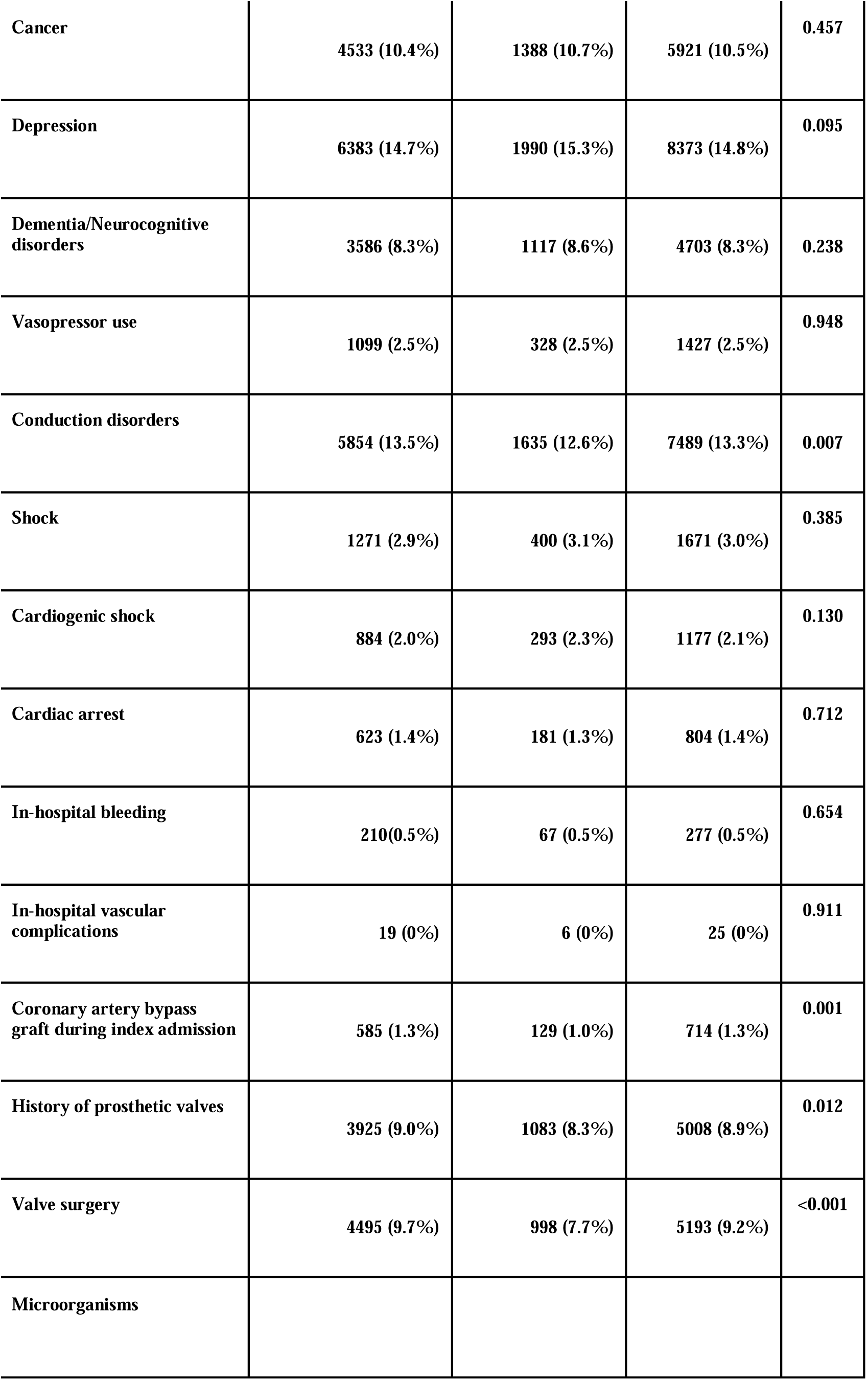
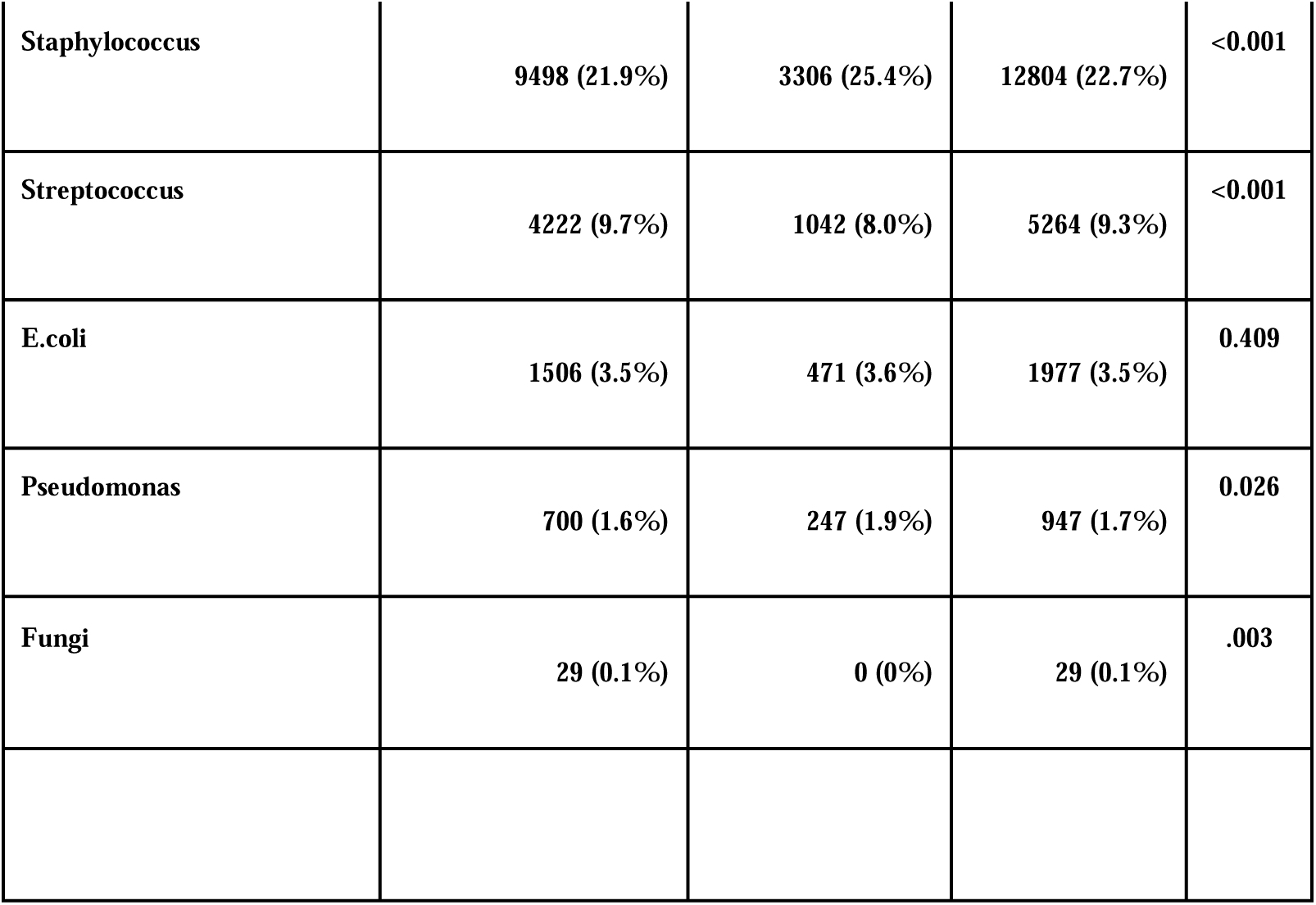
Comorbidities and procedure related factors during index admission for Infective endocarditis.

|  | **No early readmissions**<br>(n=46343; 82.2%) | **30 day readmission**<br>(n= 10048; 17.8%) | **Overall**<br>(n=56,391) | **P value** |
| --- | --- | --- | --- | --- |
| **Comorbidities** |  |  |  |  |
| **Tobacco use disorders/Smoker** | **10135 (23.4%)** | **3464 (26.6%)** | **13599 (24.1%)** | **<0.001** |
| **Alcohol use disorders** | **2103 (4.8%)** | **645 (5.0%)** | **2748 (4.9%)** | **0.597** |
| **Lipid disorders** | **15849 (36.5%)** | **4557 (35.0%)** | **20406 (36.2%)** | **0.002** |
| **Hypertension** | **11124 (25.6%)** | **2597 (20.0%)** | **13721 (24.3%)** | **<0.001** |
| **Diabetes** | **12938 (29.8%)** | **4366 (33.6%)** | **17304 (30.7%)** | **<0.001** |
| **Obesity** | **6663 (15.4%)** | **2029 (15.6%)** | **8692 (15.4%)** | **0.448** |
| **Heart failure** | **16563 (38.2%)** | **5639 (43.4%)** | **22202 (39.4%)** | **<0.001** |
| **Coronary artery disease** | **14361 (33.1%)** | **4443 (34.2%)** | **18804 (33.3%)** | **0.023** |
| **Noncardiac Chest pain** |  |  |  |  |
| **Previous Percutaneous Coronary Intervention** | **2435 (5.6%)** | **742 (5.7%)** | **3177 (5.6%)** | **0.680** |
| **Previous Coronary Artery Bypass Graft** | **229 (0.5%)** | **73 (0.6%)** | **302 (0.5%)** | **0.644** |
| **Valvular heart disease** | **11946 (27.5%)** | **3453 (26.6%)** | **15399 (27.3%)** | **0.029** |
| **Dysrhythmias** | **15136 (34.9%)** | **4710 (36.2%)** | **19846 (35.2%)** | **0.005** |
| **Cerebrovascular Accident** | **3746 (8.6%)** | **1139 (8.8%)** | **4885 (8.7%)** | **0.652** |
| **Peripheral artery disease** | **3020 (7.0%)** | **983 (7.6%)** | **4003 (7.1%)** | **0.019** |
| **Pulmonary circulatory disorders** | **4837 (11.1%)** | **1656 (12.7%)** | **6493 (11.5%)** | **<0.001** |
| **Gastrointestinal Bleed** | **1949 (4.5%)** | **431 (4.9%)** | **2580 (4.6%)** | **0.084** |
| **Chronic Obstructive Pulmonary Disease** | **7952 (18.3%)** | **2883 (22.2%)** | **10835 (19.2%)** | **<0.001** |
| **Chronic Kidney Disease** | **12567 (29.0%)** | **4709 (36.2)** | **17276 (30.6%)** | **<0.001** |
| **Acute Kidney Injury** | **13888 (32.0%)** | **4565 (35.1%)** | **18453 (32.7%)** | **<0.001** |
| **Hepatic failure** | **1232 (2.8%)** | **445 (3.4%)** | **1677 (3.0%)** | **<0.001** |
| **Thyroid disorders** | **5908 (13.6%)** | **1758 (13.5%)** | **7666 (13.6%)** | **0.781** |
| **Fluid and electrolyte disorder** | **22128 (51.0%)** | **7349 (56.5%)** | **29477 (52.3%)** | **<0.001** |
| **Acute hemorrhagic anemia** | **5249 (12.1%)** | **1455 (11.2%)** | **6704 (11.9%)** | **0.005** |
| **Coagulation disorders** | **9340 (21.5%)** | **3159 (24.3%)** | **12499 (22.2%)** | **<0.001** |
| **Cancer** | **4533 (10.4%)** | **1388 (10.7%)** | **5921 (10.5%)** | **0.457** |
| **Depression** | **6383 (14.7%)** | **1990 (15.3%)** | **8373 (14.8%)** | **0.095** |
| **Dementia/Neurocognitive disorders** | **3586 (8.3%)** | **1117 (8.6%)** | **4703 (8.3%)** | **0.238** |
| **Vasopressor use** | **1099 (2.5%)** | **328 (2.5%)** | **1427 (2.5%)** | **0.948** |
| **Conduction disorders** | **5854 (13.5%)** | **1635 (12.6%)** | **7489 (13.3%)** | **0.007** |
| **Shock** | **1271 (2.9%)** | **400 (3.1%)** | **1671 (3.0%)** | **0.385** |
| **Cardiogenic shock** | **884 (2.0%)** | **293 (2.3%)** | **1177 (2.1%)** | **0.130** |
| **Cardiac arrest** | **623 (1.4%)** | **181 (1.3%)** | **804 (1.4%)** | **0.712** |
| **In-hospital bleeding** | **210(0.5%)** | **67 (0.5%)** | **277 (0.5%)** | **0.654** |
| **In-hospital vascular complications** | **19 (0%)** | **6 (0%)** | **25 (0%)** | **0.911** |
| **Coronary artery bypass graft during index admission** | **585 (1.3%)** | **129 (1.0%)** | **714 (1.3%)** | **0.001** |
| **History of prosthetic valves** | **3925 (9.0%)** | **1083 (8.3%)** | **5008 (8.9%)** | **0.012** |
| **Valve surgery** | **4495 (9.7%)** | **998 (7.7%)** | **5193 (9.2%)** | **<0.001** |
| **Microorganisms** |  |  |  |  |
| Staphylococcus | 9498 (21.9%) | 3306 (25.4%) | 12804 (22.7%) | <0.001 |
| Streptococcus | 4222 (9.7%) | 1042 (8.0%) | 5264 (9.3%) | <0.001 |
| E.coli | 1506 (3.5%) | 471 (3.6%) | 1977 (3.5%) | 0.409 |
| Pseudomonas | 700 (1.6%) | 247 (1.9%) | 947 (1.7%) | 0.026 |
| Fungi | 29 (0.1%) | 0 (0%) | 29 (0.1%) | .003 |

The AHA study showed that the most common diagnosis was endocarditis itself, however the most common cause of readmission for infective endocarditis within 30 days in our study was a primary diagnosis of septicemia, followed by heart failure and then endocarditis^1^. Our study also found that chronic kidney disease had the highest odds of readmission compared to any other comorbidity despite only accounting for 1.9% of readmission (not even making the top 10 causes for readmission). An elevated serum creatinine in a past study was associated with higher mortality as it can reflect other issues systematically^(6)^. All of this including the results from table 5 and figure 2 shows the importance of focusing on comorbidities and appropriately managing those risk factors that may be associated with readmission, these selected variables were also chosen based on clinical importance. Considering sepsis was the most common primary diagnosis on readmission suboptimal antibiotic treatment may have been a variable involved. A study showed that although there seemed to be an increased incidence of sepsis over time, it was in fact due to increasing clinical awareness of sepsis, more screening, lower diagnostic thresholds, and better coding practices, with all of that in mind, the sepsis incidence and mortality rate has been relatively stable over the last decade^(7)^. Something to note is that treatment guidelines are not always followed in real life and therefore the index admissions are very short, and many patients are readmitted with sepsis and thus having severe financial consequences on the entire system. Additionally, a higher focus on patients with chronic kidney disease could help decrease their added risk of being readmitted as well. Outpatient parenteral antibiotic treatment (OPAT) may be the reason for the relatively short median admission time for the index hospitalization (9 days). Being able to conduct further studies to specifically track these concerns would be greatly influential. According to the new guidelines from 2015, an antibiotic treatment period of about four weeks or greater was considered appropriate^(8,3)^, changes such as a trend to stop or reduce Gentamicin use, optimization of Vancomycin troughs, recommendations to utilize Daptomycin due to being superior to Vancomycin for MSSA and MRSA were all important in improvement of management. Given the data we collected is from 2017, further trials are needed to evaluate duration of therapy and need for follow up of cultures/ inflammatory markers. In our study, Staphylococcus was the most common microorganism found in approximately 25.4% of readmissions. This could also be due to inadequate antibiotic management or increased resistance. A study also detailed how the United States has the highest incidence of infective endocarditis in the world at 15 per 100,000 people, which could be attributed to better detection and easy availability to blood cultures and echocardiography, most likely due to an increase in the aging population as well as intravenous drug abuse (staphylococcus is well known to be one of the most common microbes to be found when a patient has a history of IVDU)^(9)^.

**Table 5:**
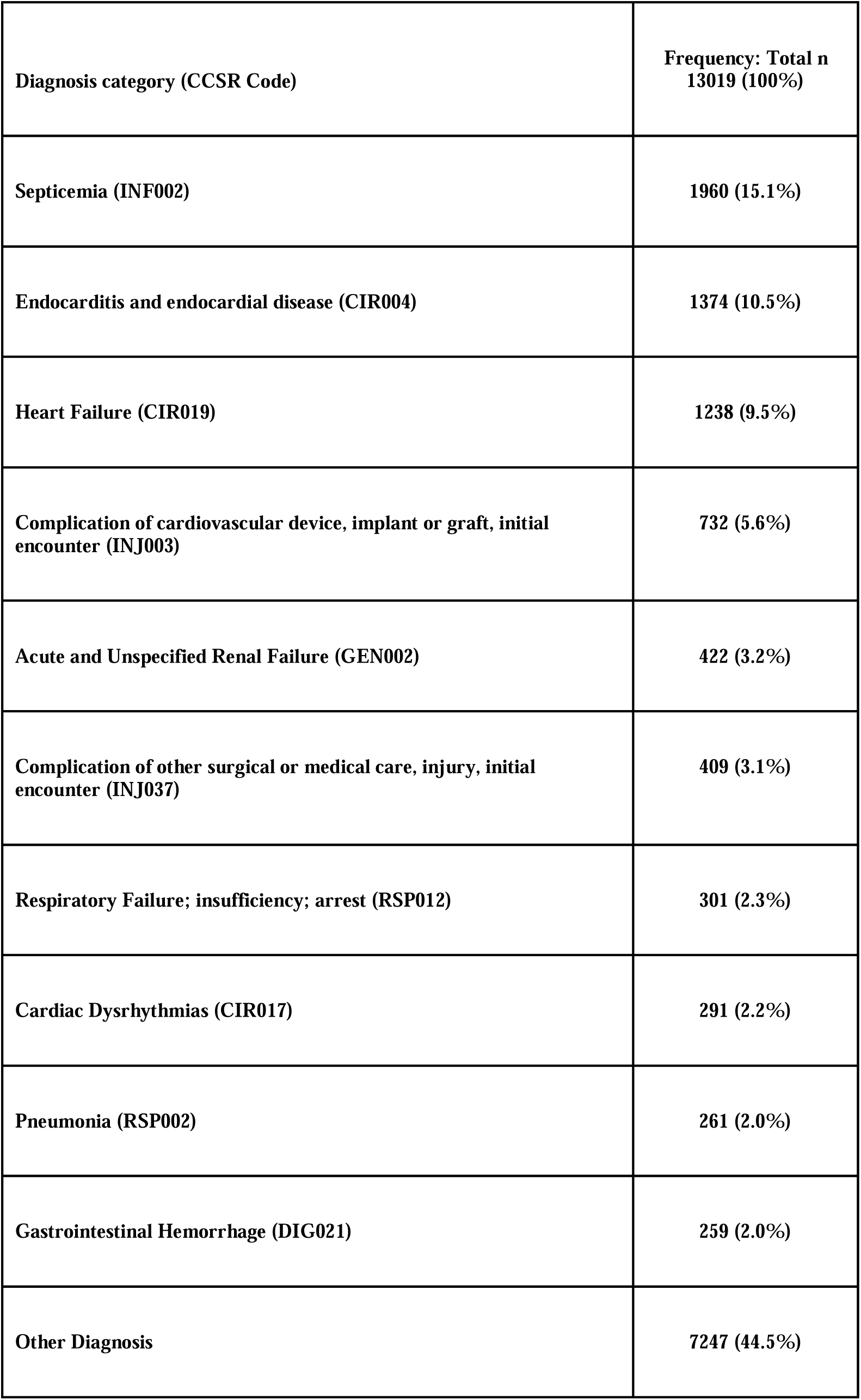
Top 10 Causes and frequencies of primary diagnosis category for readmissions encounters. (Based on the primary Clinical Classification Software refined (CCSR))

| **Diagnosis category (CCSR Code)** | **Frequency: Total n**<br>**13019 (100%)** |
| --- | --- |
| **Septicemia (INF002)** | **1960 (15.1%)** |
| **Endocarditis and endocardial disease (CIR004)** | **1374 (10.5%)** |
| **Heart Failure (CIR019)** | **1238 (9.5%)** |
| **Complication of cardiovascular device, implant or graft, initial encounter (INJ003)** | **732 (5.6%)** |
| **Acute and Unspecified Renal Failure (GEN002)** | **422 (3.2%)** |
| **Complication of other surgical or medical care, injury, initial encounter (INJ037)** | **409 (3.1%)** |
| **Respiratory Failure; insufficiency; arrest (RSP012)** | **301 (2.3%)** |
| **Cardiac Dysrhythmias (CIR017)** | **291 (2.2%)** |
| **Pneumonia (RSP002)** | **261 (2.0%)** |
| **Gastrointestinal Hemorrhage (DIG021)** | **259 (2.0%)** |
| **Other Diagnosis** | **7247 (44.5%)** |

We also aimed to find which variables had higher odds of readmission in comparison to others as shown in table 6 and figure 1. There was a variable that led to a significant decrease in odds of readmission which was valvular surgery. Even amongst readmitted patients, 7.9% of them received valve surgery. There was also an element of selection bias because surgery was obviously not an option in high risk patients even when indicated. This issue was also of concern in prior studies and considering similar results in data, there could be an argument to further weigh the risks and benefits of a more aggressive approach to valve surgery even in high risk patients. The 2015 AHA guideline promotes surgical intervention during index admission with congestive heart failure being the most common indication for ^it(1)^. Early surgery within 48 hours was compared in a recent trial to delayed surgery during the remainder of the hospital admission or follow up, which showed that those who underwent early surgery had a lower rate of death, embolic event, or reoccurrence of infective endocarditis at six months^(10)^. A recent meta-analysis also showed patients with infective endocarditis that had surgery within the first week had a lower risk of mortality^(11)^. All of this recent data suggests that there could be an argument to adjust the 2015 guidelines to push for more aggressive and earlier surgical intervention within the 1st few days of admission.

**Table 6:** Multivariable logistic regression analysis for 30 day readmissions post index admission for infective endocarditis.

|  | *P* value | Odds ratio (95% Confidence Interval) |
| --- | --- | --- |
| **Primary expected payer (Reference: Medicaid)** |  |  |
| Medicare | 0.752 | 1.01 (0.94 – 1.05) |
| Private | <0.001 | 0.73 (0.68 – 0.78) |
| Self-pay | 0.012 | 1.12 (1.02 – 1.23) |
| No charge | 0.328 | 1.00 (0.90 – 1.33) |
| Other | <0.001 | 0.61 (0.53 – 0.70) |
| **Comorbidities (Reference: no)** |  |  |
| Tobacco use disorders/Smoker | <0.001 | 1.22 (1.06–1.18) |
| Alcohol use disorders | 0.439 | 0.96 (0.87 – 1.05) |
| Lipid disorders | 0.125 | 0.96 (0.91 – 1.01) |
| Hypertension | 0.002 | 0.91 (0.86 – 0.96) |
| Diabetes | <0.001 | 1.12 (1.07 – 1.18) |
| **Obesity** | **0.070** | **0.94 (0.89 – 1.00)** |
| **Heart failure** | **<0.001** | **1.16 (1.11 – 1.22)** |
| **Coronary artery disease** | **0.192** | **1.03 (0.98 – 1.08)** |
| **Valvular heart disease** | **0.062** | **1.05 (0.99 – 1.11)** |
| **Dysrhythmias** | **<0.001** | **1.09 (1.04 – 1.14)** |
| **Previous Cerebrovascular Accident** | **0.036** | **1.07 (1.00 – 1.15)** |
| **Peripheral vascular disease** | **0.311** | **1.04 (0.96 – 1.12)** |
| **Pulmonary circulatory disorders** | **0.058** | **1.06 (0.99 – 1.13)** |
| **Gastrointestinal Bleed** | **0.498** | **1.03 (0.93 – 1.14)** |
| **Chronic Obstructive Pulmonary Disease** | **<0.001** | **1.21 (1.15 – 1.28)** |
| **Chronic Kidney Disease** | **<0.001** | **1.29 (1.22 – 1.35)** |
| **Acute Kidney Injury** | **0.264** | **1.02 (0.98 – 1.07)** |
| **Hepatic-failure** | **0.092** | **1.10 (0.98 – 1.23)** |
| **Thyroid disorders** | **0.809** | **1.00 (0.94 – 1.07)** |
| **Fluid and electrolyte disorder** | **<0.001** | **1.14 (1.09 – 1.19)** |
| **Acute hemorrhagic anemia** | **0.208** | **0.95 (0.89 – 1.02)** |
| **Coagulation disorders** | **<0.001** | **1.11 (1.06 – 1.17)** |
| **Cancer** | **0.004** | **1.10 (1.03 – 1.17)** |
| **Depression** | **0.662** | **1.01 (0.95 – 1.07)** |
| **Dementia** | **0.007** | **1.10 (1.02 – 1.19)** |
| **Valve Surgery** | **<0.001** | **0.73 (0.67 – 0.80)** |
| **Staphylococcus** | **0.003** | **1.07 (1.02 – 1.13)** |
| **Streptococcus** | **0.056** | **0.93 (0.86 – 1.00)** |
| **E. coli** | **0.638** | **1.02 (0.92 – 1.14)** |
| **Pseudomonas** | **0.158** | **1.11 (0.95 – 1.29)** |
| **History of Prosthetic Valve** | **0.034** | **0.91 (0.84 – 0.99)** |

Of note there were several comorbidities that were independent predictors of 30 day readmission after infective endocarditis. Considering the symptoms of IE can be so variable initially and clinically similar to other infectious processes, that can contribute to significant complications such as embolization and heart failure^(12,13,14)^. It is important to be able to properly differentiate all of the signs and symptoms and attribute them to the correct etiology. Other than the already mentioned CHF, diabetes mellitus was present as a comorbidity in 33.6% of readmissions (29.8% in no readmissions, P<0.001), and even more intriguing, the comorbidity with the highest percentage was 56.5% for fluid and electrolyte disorders (51% in no readmissions, P<0.001) which in itself could be due to a multitude of factors. There was a study that found hyponatremia to be the most common laboratory abnormality in patients with IE present in nearly 66% of the ICU admissions^(15)^. This data suggests that it would be critical to perform a more in-depth investigation in patients who presently have, or at risk for volatility in electrolytes and fluid balances. There was a study that argued for close interval follow up within 1, 3, 6, and 12 months post discharge^(16)^, collecting labs such as a basic metabolic panel at discharge, or even ensuring close follow-up within one week outpatient for repeat labs, may lead to improved management and a decrease in morbidity and mortality. Additionally, depression, dementia, and neurocognitive disorders did not show a significance in readmission, there has been plenty of data showing the association of mental health and prognosis. This area presents an opportunity to develop effective and efficient interventions and management techniques, that can improve and/or aid a patient’s mental status presently in order to improve adverse outcomes in the future. There was a study that summarized current evidence on psychological issues in infective endocarditis, of note it highlighted how posttraumatic stress disorder was present in 11% and which showed to have a negative impact on the quality of life of infective endocarditis patients. Overall feelings of nervousness and depressive moods were increased which effected work and activities of daily living^(17)^. There was also another study that reported to find that 1 in 4 patients had self-reported symptoms of anxiety and depression^(18)^.

Previous studies did not note tobacco use disorder and fluid and electrolyte disorders to be significance when it came to readmissions, however since the new guidelines and the latest data from 2017 we have concluded that they are indeed significant risk factors for readmission and should be critically assessed during discharge planning. Previous studies have also noted depression (increased odds of readmission) to be of significance, but we found that it did not, although it was a present as a comorbidity in 15.3% of readmissions. Finally, factors such as chronic kidney disease, COPD, hepatic failure, heart failure, coagulopathy, diabetes were all noted to be of significant risk factors in prior studies as well as ours^(1)^ The idea that these comorbidities (that in itself are associated with poor outcomes), irrespective of infectious diseases entity, show also a poor outcome in infective endocarditis, and hence, a higher likelihood for readmission is not novel idea, however we hope that by presenting more data and evidence, it can lead to more studies and further research to help reduce readmissions as it is still a problem.

While health care costs remain one of the biggest problems in the United States, an effort to decrease the financial strain that comes with readmissions should be of primary focus. A study noted that the U.S. spent approximately twice as much as other high income countries on healthcare^(19)^. The initial median admission duration from our data lasted approximately 9 days, while median readmission number was 10 days (IQR, 5-18). Essentially, we are spending more money (median, $22,059) when a patient is readmitted than on their initial admission (median, $20,241). While quick discharges have always been a motivation for hospital systems, physicians must reanalyze their assessment and plans in coherence with administration to evaluate if early discharge indeed helps financial burden overall and if early follow up may help mitigate overall costs. Additionally, a more specific and critical individualized follow up within the first few days after discharge may be considered and prediction scores be developed to help with such interventions. More work is necessary to increase awareness about infective endocarditis and factors associated with worse outcomes using social media, YouTube and Smart Phone applications as have been done for other disease conditions in the past^22, 23^.

Further research should follow on the lines of using artificial intelligence and the deep neural network to identify unidentified associations as has been pioneered inpublished studies on carotid artery stenting and Covid-19^20, 24^.

### Study limitations

By design, NRD does not allow the determination of regional variations within the dataset such that we cannot compare one state to that of another. This study does not capture data of patients who were admitted as an outpatient observation status and represents only hospital admissions. Other limitations include inaccurate coding or subjective coding by physicians as well as patients admitted outside the tracking hospitals could have been missed. As with any observational data, the results do not suggest causal relationships, as there can be other unmeasured confounders. Of note, the diagnosis codes are assigned to patients throughout an admission process based on various things such as symptoms, labs and procedures ordered, and of course results, unfortunately the NRD does not detail on validation studies to 100% confirm if a diagnosis is true, however the diagnosis codes we utilized are related to infective endocarditis and that there is a high probability that a patient actually had it. The earlier mentioned statement of the U.S having the highest incidence of IE in the world could also be due to misdiagnosis in “national database coding studies” and thus this would also be a limitation. Outpatient follow up is an extremely important component after hospital stays, but unfortunately this cannot be tracked or studied utilizing the NRD; specifically knowing how outpatient parenteral antibiotic treatment is handled is something that should be a focus in a future study.

## Conclusion

Our analysis suggests that over 1 in 5 patients admitted with infective endocarditis end up with an unplanned readmission within 30 days of discharge with the main diagnosis during readmission encounter being septicemia, heart failure, and then endocarditis itself. Surgical intervention during index admission as well as other patient and hospital related factors were associated with readmissions as well. Additionally, close follow up for sepsis and electrolyte and fluid balance disorders should be conducted to alleviate readmission burden. Patients who had co-morbidities like CKD with higher odds of readmission should have closer outpatient follow up to help prevent the readmissions as well. Data retrieved from this study that was conducted after the change in guidelines and utilization of ICD-10 may help develop protocols and management plans to reduce overall healthcare costs and improve patient outcomes.

### Perspectives

Competency in Patient Care: Patient should be counselled appropriately on risk factors that may have contributed to onset of infective endocarditis, future complications, as well as comorbidities that need to be managed much more closely that can worsen clinical conditions.

Competency in Medical Knowledge: Early IV antibiotic management is the preferred method of management for Infective Endocarditis

Interpersonal and Communication Skills: Medicine, Surgery, and Primary Care Provider need to communicate effectively with each-other as well as the patient in order to achieve the best possible outcomes at, during, and after admission and discharge.

Translational Outlook 1: Despite being a study involving data from one calendar year, readmissions from other years after the 2015 guidelines could provide more data and support for our interpretations in this study

Translational Outlook 2: There seems to be a push for surgical intervention during index admission (in patients that qualify) – there could be a more detailed comparative study done with specific surgical interventions and the times at which they were performed during an admission, i.e. within the first week of admission, vs. 2 weeks, vs. 3 weeks.

Translational Outlook 3: Closer follow-up and coordination with primary care providers outpatient should be re-evaluated in regard to repeat lab work and comorbidities that have led to a higher odds of being readmitted. A study to compare groups with closer follow-up within 1 week could be compared to those that had follow up past 1 week and those that never had follow-ups at all.

## Data Availability

All data available

## Abbreviations

NRD: Nationwide Readmission Database
IE: Infective Endocarditis
IQR: Interquartile Ratio
AHA: American Heart Association
IVDU: Intravenous Drug Use
ICD: International Classification of Diseases
CMS: Centers for Medicare & Medicaid Services
CCSR: Clinical Classification Software refined
APR-DRG: All Patient Refined-Diagnosis Related Group

## Online Table 1: ICD-10 and CCSR Codes utilized in study

| **Micro** | **Source ICD 10** | **Code (Default CCSR Cat)** |
| --- | --- | --- |
| Staphylococcus | A4101, A4102, A411, A412, B957, B958, J1520, J15211, J15212, J1529, |  |
| Streptococcus | J13, J153, A400, A401, A403, B950, B951, B953, B954, B955 |  |
| E. coli | A4151, B9620, B9629, J155 |  |
| Pseudomonas | A4152, B965, J151 |  |
| **Comorbidities** | **Source ICD 10** | **Code (Default CCSR Cat)** |
| Essential (primary) Hypertension | I10 |  |
| Hypertensive Chronic Kidney Disease with or without Heart Failure | I120, I129, I130, I1310, I1311, I132 |  |
| Renovascular Hypertension | I150 |  |
| Pulmonary Hypertension | I2723 |  |
| Post thrombotic syndrome and venous insufficiency/hypertension |  | CIR036 |
| Portal Hypertension | K766 |  |
| Heart Failure |  | CIR019 |
| Diabetes |  | END002, END003 |
| Acute Kidney Injury |  | GEN002 |
| Chronic Kidney Disease |  | GEN003 |
| Acute Myocardial Infarction |  | CIR019 |
| Nonspecific Chest Pain |  | CIR012 |
| Cardiac Dysrhythmias |  | CIR017 |
| Fluid and electrolyte disorders |  | END011 |
| Hypotension |  | CIR031 |
| Complication of other surgical or medical care, injury, initial encounter |  | INJ037 |
| Urinary Tract Infection |  | GEN004 |
| Tobacco Use Disorder |  | MBD024 |
| Alcohol Use Disorder |  | MBD017 |
| Obesity |  | END009 |
| Depression |  | MBD002 |
| Heart Valve Procedures | CCS Category 43, Multi CCS LVL1 7 |  |

## CENTRAL ILLUSTRATION

